# Hospital-Acquired Acute Kidney Injury in Non-critical Medical Patients in a Developing Country Tertiary Hospital: Incidence and Predictors

**DOI:** 10.1101/2023.09.21.23295890

**Authors:** Nahom Dessalegn Mekonnen, Tigist Workneh Leulseged, Buure Ayderuss Hassen, Kidus Haile Yemaneberhan, Helen Surafeal Berhe, Nebiat Adane Mera, Anteneh Abera Beyene, Lidiya Zenebe Getachew, Birukti Gebreyohannes Habtezgi, Feven Negasi Abriha

**Author notes:** Corresponding author: Tigist Workneh Leulseged.

## Abstract

**Background:** Acute kidney injury (AKI) is a frequent complication in critical patients leading to worse prognosis. Although the consequences of AKI are worse among critical patients, AKI is also associated with less favorable outcomes in non-critical patients. Hence, understanding the magnitude of the problem in these patients is crucial, yet there is a scarcity of evidence in non-critical settings, especially in resource limited countries. Hence, the study aimed at determining the incidence and predictors of hospital acquired acute kidney injury (HAAKI) in non-critical medical patients who were admitted at a large tertiary hospital in Ethiopia.

**Methods:** A retrospective chart review study was conducted among 232 hospitalized non-critical medical patients admitted to St. Paul’s Hospital Millennium Medical College between January 2020 and January 2022. Data was characterized using frequency and median with interquartile range. To identify predictors of HAAKI, a log binomial regression model was fitted at a p value of ≤ 0.05. The magnitude of association was measured using adjusted relative risk (ARR) with its 95% CI.

**Results:** During the median follow-up duration of 11 days (IQR, 6-19 days), the incidence of HAAKI was estimated to be 6.0 per 100 person-day observation (95% CI= 5.5 to 7.2). Significant predictors of HAAKI were found to be having type 2 diabetes mellitus (ARR=2.36, 95% CI= 1.03, 5.39, p-value=0.042), and taking vancomycin (ARR=3.04, 95% CI= 1.38, 6.72, p-value=0.006) and proton pump inhibitors (ARR=3.80, 95% CI = 1.34,10.82, p-value=0.012).

**Conclusions:** HAAKI is a common complication in hospitalized non-critical medical patients, and is associated with a common medical condition and commonly prescribed medications. Therefore, it is important to remain vigilant in the prevention and timely identification of these cases and to establish a system of rational prescribing habits.

## BACKGROUND

Acute kidney injury (AKI) is defined as a sudden decrease in renal function that can result in an accumulation of toxins in the blood (1-2). Depending on where it occurs, AKI is classified as either community-acquired AKI or hospital-acquired AKI (HAAKI). With early diagnosis and management, AKI is fully reversible in both cases, with no short-or long-term complications. HAAKI is a common complication in hospitalized patients, affecting millions of people each year. Unlike community-acquired AKI, HAAKI can often be missed, especially in non-critical settings and hence can lead to an increased risk of complications and mortality. HAAKI is also associated with increased healthcare costs, imposing an additional burden on the healthcare system (3-9).

The incidence of HAAKI is reported to be high, affecting almost a quarter of hospitalized patients globally. However, the overall incidence of HAAKI, especially severe cases, is significantly higher in sub-Saharan Africa, where over five times as many patients present with advanced stage as compared to other settings. This in turn is associated with a greater risk of complications and mortality (10-17). However, the majority of these findings are based on research that are conducted among cases with community-acquired AKI or HAAKI in critical patients, limiting our understanding of HAAKI in non-critical patients. Only a few studies have examined HAAKI in a non-critical setting and found that it is also a prevalent and serious complication that requires attention. These studies additionally revealed that exposure to nephrotoxic medications is the most common cause of HAAKI in non-critical medical patients, as opposed to surgical patients and those in the ICU, where sepsis is the most common cause. This implies that HAAKI has different predictors in different settings, and that we should use this knowledge to guide screening and more effectively identify patients at risk (18-24).

Evidence gaps in non-critical patients, especially in Africa, have led to poor early screening and diagnosis of HAAKI, resulting in late diagnosis and serious consequences for patients and the healthcare system. As a result, prompt diagnosis and management of AKI in hospitalized patients is critical in any care setting, particularly in countries where providing standard of care can be hampered due to inadequate health care infrastructure and where prevention is of paramount importance (18,19, 25). Hence, the study aimed at determining the incidence and predictors of HAAKI in non-critical medical patients who were admitted to St. Paul’s Hospital Millennium Medical College between January 2020 and January 2022 in Ethiopia.

## METHODS

### Study Setting and Design

An institution-based retrospective chart review study was conducted among non-critical medical patients who were admitted to one of the largest tertiary hospitals in Ethiopia, St. Paul’s Hospital Millennium Medical College (SPHMMC). SPHMMC is one of Ethiopia’s largest tertiary referral hospitals, located in the country’s capital, Addis Ababa. It is also home to the country’s only renal transplant center and has a well-equipped dialysis unit that serves as the primary national referral center for cases that require dialysis.

### Population and Sample Size

Eligible participants who were managed at the hospital during the two years observation period were identified to be included in the study based on the following criteria; patients who had no acute kidney injury or underlying chronic renal condition at the time of admission, who were admitted for at least 48 hours, and completed a follow-up at the hospital with the major exposures and outcome recorded on their medical charts. Patients who were transferred to the ward from any critical ward or intensive care units were further excluded. Finally, a total of 232 eligible cases were included in the final analysis.

**Figure 1:**
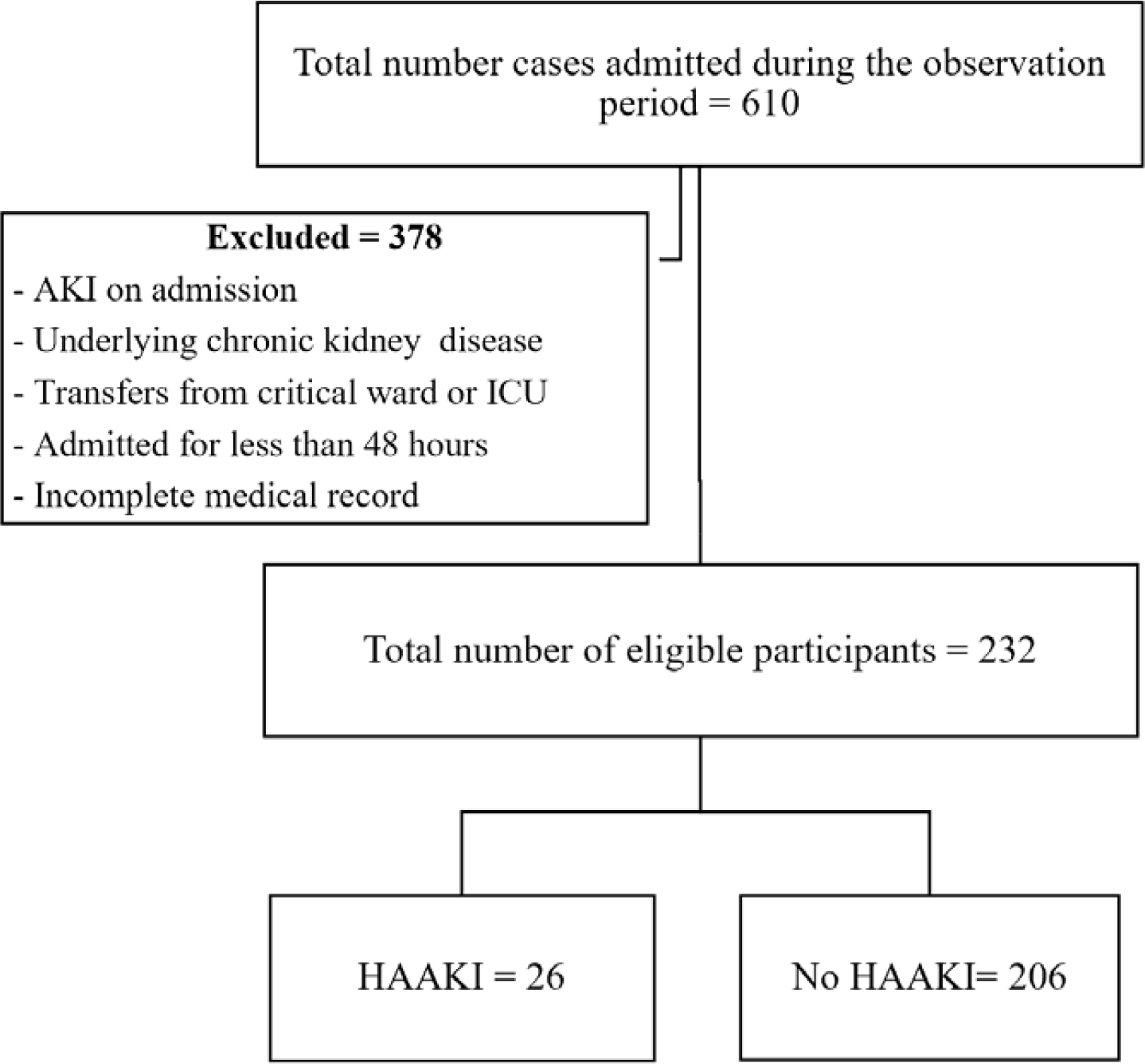
Flow chart showing the selection of study participants to be included in the final, January 2020 to January 2022, Ethiopia

Since the total number of eligible participants identified were small, all of them were included in the study, and the study’s achieved power was determined post-hoc using G*Power 3.19.4. Accordingly, using a two-tailed z-test at a 5% level of significance and the difference in proportion of HAAKI between groups based on the identified significant exposures (sepsis, vancomycin, and PPIs), the highest achieved power of the study was found to be 88.2%.

### Operational Definition

#### Hospital Acquired Acute Kidney Injury (HAAKI)

an increase in serum creatinine by ≥ 0.3mg/dl within 48 hours or increase in serum creatinine to ≥ 1.5 times baseline, which is known or presumed to have occurred within the previous 7 days (26).

- **Stage 1 AKI:** an increase in serum creatinine to 1.5 to 1.9 times baseline, or increase in serum creatinine by ≥0.3 mg/dL, or reduction in urine output to <0.5 mL/kg/hour for 6 to 12 hours.
- **Stage 2 AKI:** an increase in serum creatinine to 2.0 to 2.9 times baseline, or reduction in urine output to <0.5 mL/kg/hour for ≥12 hours.
- **Stage 3 AKI:** an increase in serum creatinine to 3.0 times baseline, or increase in serum creatinine to ≥4.0 mg/dL or reduction in urine output to <0.3 mL/kg/hour for ≥24 hours, or anuria for ≥12 hours, or the initiation of kidney replacement therapy.

## Data Collection Procedures and Quality Assurance

A pretested data abstraction tool, using Google Form, was used to extract data from the medical records of the enrolled participants. Data on baseline sociodemographic and clinical parameters were collected by three trained General Practitioners with the supervision of a senior internal medicine resident. The electronic data was exported into STATA software version 17.0 (College Station, TX) for data management and analysis. Before beginning the data analysis step, data management was completed, which included data cleaning, transformation, cross-referencing inconsistencies, and imputations.

## Statistical Analysis

The characteristics of the study participants were summarized using a simple count with percentage and a median with interquartile range. The incidence of HAAKI was estimated using incidence density with its 95% CI by taking the number of new cases of HAAKI and the total person day (PD) observation of the study participants.

A log binomial regression model was used to identify significant predictors of HAAKI. First, a univariate analysis was performed at a significance level of 25%. The variables that passed the univariate analysis selection criteria and those that were considered to be clinically relevant were then fitted into the final multivariable regression model. The final model’s adequacy was evaluated using the goodness of fit test (Pearson Chi-Square = 0.962) and the omnibus test (p-value = 0.006), both of which demonstrated that the data fitted the model adequately. From the final model, at a p value of ≤ 0.05, the magnitude of association between the significant predictors and development of HAAKI was measured using adjusted relative risk (ARR) with its 95% CI.

## RESULTS

### Baseline Socio-demographic and Clinical Characteristics

The majority of the patients were younger than 60 years (70.7%) and were males (55.6%). Hypertension was the most frequently identified preexisting chronic medical illness recorded in 89 (38.4%) patients, followed by type 2 diabetes mellitus (T2DM) in 40 (17.2%) and chronic lung disease in 23 (9.9%) patients. Pneumonia and heart failure were the most common primary admission diagnosis in 90 (38.8%) and 62 (26.7%) patients, respectively.

On admission, more than three-quarters were started on cephalosporin (77.6%) and more than half were taking anticoagulants (57.3%) and proton pump inhibitors (PPIs) (56.9%). Furthermore, over a quarter were taking vancomycin (39.2%), diuretics (30.6%), and steroids (27.2%). The results of the blood tests revealed that 58 (25.0%) had leukocytosis, 91 (39.2%) were anemic, 68 (29.3%) had hyponatremia, and 37 (15.9%) had hypokalemia. **(Table 1)**

**Table 1:** Baseline socio-demographic and clinical characteristics among non-critical medical patients at SPHMMC in Ethiopia, January 2020 to January 2022 (n=232)

| Variable | Frequency (%) | Variable | Frequency (%) |
| --- | --- | --- | --- |
| **Age group (in years)** |  | **Medication** |  |
| <40 | 90 (38.8) | Cephalosporin | 180 (77.6) |
| 40-59 | 74 (31.9) | Anticoagulants | 133 (57.3) |
| ≥ 60 | 68 (29.3) | PPIs | 132 (56.9) |
| **Sex** |  | Vancomycin | 91 (39.2) |
| Male | 129 (55.6) | Diuretics | 71 (30.6) |
| Female | 103 (44.4) | Steroids | 63 (27.2) |
| **Comorbidity** |  | ACEI/ARBs | 41 (17.7) |
| Hypertension | 89 (38.4) | Macrolides | 38 (16.4) |
| DM | 40 (17.2) | Anti TB | 23 (9.9) |
| Chronic lung disease | 23 (9.9) | Antiplatelet | 17 (7.3) |
| Cardiovascular disease | 15 (6.5) | Penicillin | 14 (6.0) |
| HIV | 13 (5.6) | Quinolones | 6 (2.6) |
| Malignancy | 9 (3.9) | **Laboratory parameters** |  |
| **Admission diagnosis** |  | Normal WBC | 150 (64.7) |
| Pneumonia | 90 (38.8) | Low WBC | 24 (10.3) |
| Heart failure | 62 (26.7) | High WBC | 58 (25.0) |
| DVT/PE | 41 (17.7) | Normal Hemoglobin | 109 (47.0) |
| Stroke | 28 (12.1) | Low Hemoglobin | 91 (39.2) |
| Sepsis | 23 (9.9) | High Hemoglobin | 32 (13.8) |
| TB | 21 (9.1) | Normal Sodium | 155 (66.8) |
| CNS infection | 18 (7.8) | Low Sodium | 68 (29.3) |
| Hepatitis | 15 (6.5) | High Sodium | 9 (3.9) |
| GI infection | 11 (4.7) | Normal Potassium | 184 (79.3) |
| COPD | 9 (3.9) | Low Potassium | 37 (15.9) |
| Upper GI Bleeding | 8 (3.4) | High Potassium | 11 (4.7) |

### Incidence of HAAKI

The study participants were observed for a median of 11 days (IQR, 6-19 days). During this time, 26 of the 232 hospitalized patients developed HAAKI, resulting in an incidence density of 6.0 per 100 PD observation (95% CI= 5.5 to 7.2). From the 26 patients with HAAKI, 15 had stage 1 AKI, 7 had stage 2 AKI, and 4 had stage 3 AKI, and none required dialysis.

### Predictors of HAAKI

To identify the predictors of HAAKI, a multivariable log binomial regression analysis was run at 5% level of significance. Age group, sex, hypertension, T2DM, sepsis, DVT/PE, macrolides, vancomycin, ACEIs/ARBs, PPIs, anticoagulants, steroids, WBC, hemoglobin, and sodium level were fitted in the model. Consequently, from the final model, T2DM, vancomycin and PPIs were found to be the only significant predictors of HAAKI development. Patients with T2DM were 2.36 times more likely to develop HAAKI than those with no T2DM (**ARR=2.36, 95% CI= 1.03, 5.39, p-value=0.042)**. In addition, the risk of developing HAAKI was increased by3.04 times among patients taking vancomycin **(ARR=3.04, 95% CI= 1.38, 6.72, p-value=0.006)** and by 3.80 times among patients taking PPIs **(ARR=3.80, 95% CI = 1.34,10.82, p-value=0.012)** compared to those who were not prescribed these medications throughout their stay. (**Table 2**)

**Table 2:** Predictors of HAAKI among non-critical medical patients at SPHMMC in Ethiopia, January 2020 to January 2022 (n=232)

| Variable |  | HAAKI |  | CRR (95% CI) | ARR (95% CI) | p-value |
| --- | --- | --- | --- | --- | --- | --- |
|  |  | Yes (n=26) | No (n=206) |  |  |  |
| Age group (in years) | <40 | 5 | 85 | 1 | 1 |  |
|  | 40-59 | 10 | 64 | 2.43 (0.87, 6.80) | 1.79 (0.57, 5.57) | 0.317 |
|  | >=60 | 11 | 57 | 2.91 (1.06, 7.99) | 2.39 (0.74, 7.7.4) | 0.146 |
| Sex | Male | 17 | 112 | 1 | 1 |  |
|  | Female | 9 | 94 | 0.66 (0.31, 1.43) | 0.57 (0.25, 1.28) | 0.173 |
| Hypertension | No | 12 | 131 | 1 | 1 |  |
|  | Yes | 14 | 75 | 1.88 (0.91, 3.87) | 1.58 (0.56, 4.41) | 0.386 |
| T2DM | No | 18 | 174 | 1 | 1 |  |
|  | Yes | 8 | 32 | 2.13 (0.99, 4.56) | **2.36 (1.03, 5.39)** | **0.042\*** |
| Sepsis | No | 22 | 187 | 1 | 1 |  |
|  | Yes | 4 | 19 | 1.65 (0.62, 4.38) | 1.75 (0.59, 5.16) | 0.308 |
| DVT/PE | No | 21 | 170 | 1 | 1 |  |
|  | Yes | 5 | 36 | 1.11 (0.44, 2.77) | 1.68 (0.59, 4.78) | 0.335 |
| Macrolides | No | 21 | 173 | 1 | 1 |  |
|  | Yes | 5 | 33 | 1.22 (0.49, 3.02) | 1.76 (0.65, 4.78) | 0.265 |
| Vancomycin | No | 10 | 131 | 1 | 1 |  |
|  | Yes | 16 | 75 | 2.48 (1.18, 5.22) | **3.04 (1.38, 6.72)** | **0.006\*** |
| ACEI/ARBs | No | 21 | 170 | 1 | 1 |  |
|  | Yes | 5 | 36 | 1.11 (0.44, 2.77) | 0.46 (0.16, 1.33) | 0.152 |
| PPIs | No | 4 | 96 | 1 | 1 |  |
|  | Yes | 22 | 110 | 4.17 (1.48, 11.71) | **3.80 (1.34, 10.82)** | **0.012\*** |
| Anticoagulants | No | 8 | 91 | 1 | 1 |  |
|  | Yes | 18 | 115 | 1.68 (0.76, 3.69) | 1.67 (0.72, 3.86) | 0.232 |
| Steroids | No | 21 | 148 | 1 | 1 |  |
|  | Yes | 5 | 58 | 0.64 (0.25, 1.62) | 0.55 (0.21, 1.42) | 0.216 |
| WBC | Normal | 17 | 133 | 1 | 1 |  |
|  | Low/High | 9 | 73 | 0.97 (0.45, 2.08) | 0.71 (0.33, 1.53) | 0.386 |
| Hemoglobin | Normal | 11 | 98 | 1 | 1 |  |
|  | Low/High | 15 | 108 | 1.21 (0.58, 2.52) | 1.08 (0.51, 2.32) | 0.839 |
| Sodium | Normal | 14 | 141 | 1 | 1 |  |
|  | Low/High | 12 | 65 | 1.73 (0.84, 3.55) | 1.73 (0.73, 4.07) | 0.211 |

CRR= Crude Relative Risk, ARR= Adjusted Relative Risk, \* = statistically significant

## DISCUSSION

The current study was conducted with the aim of determining the incidence and predictors of HAAKI in non-critical medical patients who were admitted to St. Paul’s Hospital Millennium Medical College between January 2020 and January 2022 in Ethiopia. The study included 232 eligible participants, the majority of whom were younger than 60 years (70.7%), had hypertension (38.4%), were admitted primarily for pneumonia (38.8%), and were taking cephalosporin (77.6%), anti-coagulants (57.3%) and proton pump inhibitors (PPIs) (56.9%).

The incidence of HAAKI was determined to be 6.0 per 100 PD observations. Although it is not as incidental as in a critical care setting, this is a clinically significant rate considering that it is conducted in non-critical patients, indicating that HAAKI is not uncommon in this population as well. However, there is still a potential that this is an underestimation of the true magnitude of the problem, as creatinine levels are not routinely assessed in non-critical patients in the study setting unless they have a known underlying renal condition or show deterioration in their medical condition, resulting in missing of cases that revert without complication.

The significant predictors of development of HAKKI in a non-critical setting were identified to be having T2DM, taking vancomycin and PPIs. Patients with T2DM were 2.36 times more likely to develop HAAKI than those with no T2DM. Diabetes, especially in the uncontrolled state, is one of the well-known risk factors for kidney injury, particularly in a hospital setting due to exposure to a multitude of additional factors that might compound the risk of damage. Previous studies also support this finding (23,24, 27-29).

Moreover, patients taking vancomycin plus PPIs were more than three times as likely to develop HAAKI. Vancomycin and PPIs are both reported to be nephrotoxic. Studies have shown that vancomycin causes acute kidney injury through several mechanisms, the most well-known of which being oxidative stress in the kidney’s proximal tubules. PPIs also increase the risk of acute interstitial nephritis; the specific mechanism is unknown, although it is thought to be linked to immune-mediated responses, oxidative stress, and possibly renal tubular death (30, 31). The nephrotoxic effect of these drugs is especially exacerbated when they are administered frequently in large doses to patients who have underlying medical conditions and complications such as dehydration and sepsis. Nephrotoxic drugs are the most common risk factors for HAAKI in medical patients, unlike surgical patients as reported in another study as well (18).

In summary, this study provides valuable clinical insights into the incidence and predictors of HAAKI in non-critical medical patients in a hospital with a nephrology unit that functions relatively well in a developing country and hence the findings can be applicable to comparable settings. However, the effect of some additional behavioral, clinical factors and medication related factors, including dose-response relationships and potential drug-drug interactions, that can contribute to HAAKI development are not studied as these factors were not recorded consistently.

## CONCLUSION

HAAKI is found to be a common finding in hospitalized non-critical medical patients implying the need to stay vigilant in timely identification and management of these cases in order to prevent further complication and mortality. And it is significantly associated with T2DM, a common chronic medical condition, and taking vancomycin and PPI, commonly prescribed medications, indicating that a higher proportion of hospitalized patients are at risk of HAAKI. Hence, there is a need to establish a system to timely diagnose AKI and to practice rational prescription habits to minimize risk of exposure, whenever possible, as well as to carefully monitor patients while they are on these treatments.

## Data Availability

All relevant data are available upon reasonable request from the corresponding author.

## Declaration

### Ethics approval and consent to participate

The study was carried out after obtaining ethical approval from the institutional review board of St. Paul’s Hospital Millennium Medical College (SPHMMC-IRB) (Ref. No. PM23/385) for the conduct of the study using secondary data. Anonymity of the participants was maintained by use of medical record number in the research report. No other personal identifiers of the patients were used in the research report. Access to the collected information was limited to the investigators and confidentiality was maintained throughout the project.

### Consent for publication

Not applicable

### Availability of data and materials

All relevant data are available upon reasonable request from the corresponding author.

### Authors’ Contribution

NDM and TWL conceived and designed the study. NAM, HSB, AAB, KHY, BAH, FNA, LZG, and BGH contributed to the conception and design of the study. NDM and TWL performed statistical analysis, and drafted the initial manuscript. NAM, HSB, AAB, and BAH contributed to the statistical analysis and interpretation of the findings. KHY, FNA, LZG, and BGH revised the manuscript. All authors approved the final version of the manuscript.

## Acknowledgment

The authors would like to thank all individuals involved in the data collection, supervision, and facilitation of the research work.

